# Stigma among Ebola Disease Survivors in Mubende and Kassanda districts, Central Uganda, 2022

**DOI:** 10.1101/2024.05.07.24307005

**Authors:** Gorreti Marie Zalwango, Sarah Paige, Richard Migisha, Brenda Nakafeero Simbwa, Edirisa Junior Nsubuga, Alice Asio, Zainah Kabami, Jane Frances Zalwango, Peter Chris Kawungezi, Mercy Wendy Wanyana, Patrick King, Hellen Nelly Naiga, Brian Agaba, Robert Zavuga, Giulia Earle-Richardson, Benon Kwesiga, Lilian Bulage, Daniel Kadobera, Alex Riolexus Ario, Julie R. Harris

## Abstract

**Background:** Ebola disease survivors often experience stigma in multiple forms, including felt (perceived) stigma, enacted (action-based) stigma, and structural (institutional) stigma. On September 20, 2022, Uganda declared a Sudan Virus Disease (SVD, caused by *Sudan ebolavirus*) outbreak after a patient with confirmed Sudan virus (SUDV) infection was identified in Mubende District. The outbreak led to 142 confirmed and 22 probable cases over the next two months. We examined the types of stigma experienced by survivors and their household members and its effect on their well-being.

**Methods:** We conducted a qualitative study during January 2023 in Mubende and Kassanda Districts. We conducted in-depth and key informant interviews with ten SVD survivors, ten household members of SVD survivors, and ten key informants (district officials and health workers in the affected communities). Interviews were recorded, translated, transcribed, and analyzed thematically.

**Results:** Survivors reported experiencing isolation and rejection by community members and loss of work. They reported being denied purchases at shops or having their money collected in a basket and disinfected (enacted stigma), which led to self-isolation (felt stigma). Educational institutions denied admission to some students from affected homes, while parents of children in some affected families stopped sending children to school due to verbal abuse from students and teachers (structural stigma). Prolonged SVD symptoms and additional attention to survivors from responders (including home visits by health workers, public distribution of support items, and conspicuous transport from home to the survivor’s clinic) were perceived as aggravating both felt and enacted stigma. Even after the outbreak had been declared over, survivors felt that they were still considered a threat to the community.

**Conclusion:** Survivors experienced felt stigma, enacted stigma, and structural stigma. Strengthening community engagement to counteract stigma, rethinking response activities that aggravate stigma, management of long-term SVD symptoms for survivors, integrated response interventions by partners, private distribution of support items, and increasing awareness and sensitization through video messages could reduce stigma among persons affected in future similar outbreaks.

## Introduction

Ebola disease (EBOD) is a severe and frequently lethal disease caused by infection with an ebolavirus. EBOD outbreaks typically start following exposure of a human to the body fluids of an infected bat or primate, followed by human-to-human transmission through contact with infected bodily fluids or contaminated fomites [1]. There are six species of Ebola virus: *Bundibugyo ebolavirus, Zaire ebolavirus, Sudan virus, Reston ebolavirus, Bombali ebolavirus*, and *Taï Forest ebolavirus*. Bundibugyo ebolavirus, Zaire ebolavirus, and Sudan virus have caused epidemics in Africa, with case-fatality rates ranging from 25-90% [2, 3].

Several EBOD outbreaks have been registered in Africa in the last three decades [4]. Uganda has registered five EBOD outbreaks since 2000 [5, 6]. On September 20, 2022, Uganda declared the sixth EBOD outbreak in the country, caused by Sudan virus (SUDV). Sudan virus disease (SVD) has been the most frequent EBOD in Uganda[7]. The outbreak led to 142 confirmed and 22 probable cases; 87 patients survived [8]. While nine districts had at least one case, two districts - Mubende and Kassanda – had the vast majority of cases [8].

Survivors of the infection often experience stigma and mental health challenges associated with being stigmatized [9-13]. Stigma around EBOD survivors is primarily associated with fear of ongoing contagion, and has led to evictions, intimate partnership dissolution, termination of employment, abandonment, and physical violence [14]. Stigma may take three forms: enacted (when survivors are discriminated against by community members), felt (when the stigmatized persons endorse the negative views from others), and structural (perpetuated by institutions such as medical, educational, and employment institutions) [15].

Stigma can affect the willingness of affected persons to seek care and the community’s acceptance of prevention and case management packages [6]. As a result, addressing disease-associated stigma starting early in an epidemic through comprehensive medical and psychosocial support and community interventions can be critical. Support provided to survivors after discharge has been linked to better coping, accelerated recovery, and faster restoration of their sense of dignity [16]. Furthermore, community efforts, such as training survivors as peer health promoters to support disease prevention and response efforts, may also help in the psychosocial recovery of individuals and in their de-stigmatization within communities [17].

Anticipating stigmatization of survivors after the outbreak, multiple interventions were provided by Ministry of Health and partners to reduce stigma and quicken reintegration of survivors in the communities. Starting in November 2022, the Ministry of Health, supported by USAID, CDC, and other partners, established a national Ebola survivors program to support the 87 survivors of the 2022 Uganda outbreak [18]. Support activities included community dialogue meetings during which survivors, their families, and communities received information about SUDV and how to help the survivors continue to recover. In addition, response staff traveled to communities to conduct health education using loudspeakers. Survivors were provided with 400,000 Uganda shillings (approximately $100 USD) each from the Bangladesh Rural Advancement Committee (BRAC) Uganda to help compensate for time lost at work. They further received in-kind support such as mattresses, clothes, and food from UNICEF and Save the Children. These supplies were issued publicly at the subcounty headquarters during November 2022 to January 2023 [19]. The survivors’ clinic provided a branded van to collect survivors from the community and bring them in for biweekly physical checkups and psychosocial support; survivors’ clinic staff and other health partners also provided psychosocial support to survivors and their family members during visits to the affected communities. Despite these efforts, stigma may continue to negatively affect EBOD survivors. To better understand how to promote the well-being of survivors in Uganda, we examined the stigma experienced by EBOD survivors and their household members, the aggravating factors to stigma and how it affected their lives to inform stigma control measures for improved epidemic response and survivor support in future outbreaks. We further explored opportunities for reducing stigma through key informant interviews conducted among local government leaders on stigma reduction recommendations.

## Methods

### Study design and setting

We conducted a qualitative study to explore the perceptions and experiences of SVD survivors and their household members using in-depth interviews of survivors, household members, and local government officials with knowledge of the issue (“key informants”). We focused on Mubende and Kassanda districts since these were the most affected areas and had a large number of SVD survivors. Neither of these districts had experienced an EBOD outbreak before.

### Study population

Participants for this study were SVD survivors in the Mubende and Kassanda districts and members of the same household as the survivors aged 18 years or above. However, an exception was made to include emancipated minors (children above 16 years living independently). In these two regions, there are a total of 63 known survivors [40 (63%) men, 23 (37%) women and 9 (14%) children <18 years]. Of the 63 survivors, 36 (57%) were from Mubende District. Children living with their parents or guardians and individuals whose physical and psychological health limited them from providing information through the interview process were excluded from this study due to limited resources. We identified survivors using a discharge list from the Ebola treatment unit (ETU) and located them in the communities with the help of community health workers (CHW). All participants provided written informed consent to participate.

We recruited survivors and household members from each district for in-depth interviews (IDI) based on their availability at home during data collection activities. Only one survivor or household member was recruited per household for IDIs to increase the variability of findings. We also engaged local government leaders from each district as key informants on how stigma could be reduced for improved wellbeing of the SVD survivors. These included health workers attached to health facilities in SVD-affected communities, the District Health Officer (DHO), Resident District Commissioner (RDC), and the District Surveillance focal person (DSFP).

Consistent with best practices, recruitment and interviewing continued until all information received from participants repeated what had been previously collected from other participants (“saturation”)[20]. Saturation was reached with 20 in-depth interviews (10 survivors and 10 household members) and 10 key informant interviews.

### In-depth interviews

Using an interview guide that provided open-ended questions and discussion prompts related to survivor feelings and experiences, the authors (GMZ and BS) conducted in-depth interviews in the local language (Luganda) with the SVD survivors and their household members using the interview guide in the appendix. Areas explored included: experiences of stigma, possible stigma instigating factors or actions, how stigma affected their lives, possible suggestions for the control of stigma, and any additional support required. In addition, we collected data on age, sex, place of residence, SVD status (survivor or household member), number of SVD patients in household, and SVD deaths in household from survivors and their household members.

Key informant interviews (also using an interview guide) explored community perceptions and actions towards survivors and their household members, health concerns of survivors, and recommendations for improvement. Information from the interviews was recorded in electronic form using audio digital recorders. Data were collected from participants in both districts concurrently until saturation was attained [21].

### Qualitative analysis approach

Researchers used a thematic analysis approach, in which text is initially coded, then themes are created, using common codings to “build” themes that describe perspectives and experiences related to the topic of interest. Thematic analysis is based in phenomenology [22], an approach which seeks to understand how people *experience* and *describe* their own situation. However, in recent years thematic analysis has become a research approach in its own right [23]. Analysis was conducted using the CDC Excel Tool for Thematic Analysis (version 2.0), which provides both a theory-based analytical framework and a practical tool for completing the analysis.

### Data analysis

Recordings were translated to English and transcribed at the end of each data collection day and stored by the principal investigator in a password protected computer. Participants were given an identification number based on the type of interview and order in which they were interviewed.

After data collection, transcripts were reviewed by the authors (GMZ and BS), coded and analyzed thematically using the CDC Excel Tool for Thematic Analysis V2.0 (10.18.22) (“Excel Tool”) to bring out the story of lived experiences and recommendations. The same coding scheme was used for all the 30 transcripts at the same time (Appendix 2). The initial coding was done by the corresponding author and later verified by 2 other authors to ensure that all relevant statements to the objectives under study were accommodated in the existing codes.

We reviewed the common codes and created statements about stigma that one or more common codes suggested. Once all of the texts were fully coded in the CDC tool, researchers created simple tables showing the frequency distributions of the codings, with the most frequently used codes at the top, and the least frequently used codes at the bottom. This provided a first look at common codes to consider for development into themes. This also allowed researchers to doublecheck coding and decide if any revisions to the coding scheme were needed. Using the coded text, we developed themes around stigma among SVD survivors.

## Results

### Survivors and families interviewed

We interviewed 10 (16%) of the 63 survivors in Mubende and Kassanda districts. All survivors approached consented to participate. Of the 10 survivors interviewed, six (60%) were female. Median age was 33 years (range: 18–70 years). We further interviewed 10 members of survivor households that were not sampled for survivors; two additional household members declined to consent. Eight (80%) household members were females; all eight were mothers or spouses to the survivors and two (20%) were siblings. The median age of household members was 32 years (range: 19–57 years). Of the 10 key informants, seven (70%) were male and the median was 37 years (range: 28–58 years).

### Text coding results

Altogether, the 30 interview transcripts yielded fourteen (14) codings. **Table 1** shows the frequency of the codes used. Under the ‘types of stigma’ theme, enacted stigma was the most frequently reported (24 coded segments, 55% of all codings under this theme) and institutional stigma was the least reported (eight coded segments, 18% of all codings under this theme). The ‘causes of stigma’ theme yielded 4 subthemes, with public monetary and in-kind support being the most frequently reported (26 coded segments, 34% of all codings in this theme) followed by extra attention to survivors (24 coded segments, 31% of all codings in this theme). The most frequent ‘effect of stigma’ subtheme was economic (22 coded segments, 55% of all codings in this theme), while the least frequent was social effects (six coded segments, 15% of all codings in this theme).

**Table 1:** Frequency of codes used following analysis of data on stigma among Sudan virus disease survivors and their household members in Mubende and Kassanda districts, Uganda, 2022.

| **Themes and sub-themes** | **Count of transcript segments coded** |
| --- | --- |
| **Types of stigma** | **44** |
| Enacted stigma | 24 |
| Felt stigma | 12 |
| Institutional stigma | 8 |
| **Causes of stigma** | **77** |
| Fear that survivors who looked unwell were still ill | 13 |
| The extra attention to survivors | 24 |
| Public monetary and in-kind support | 26 |
| Risk communication about survivors was confusing | 14 |
| **Effects of stigma** | **40** |
| Economic effects | 22 |
| Social effects | 6 |
| Education disruption | 12 |
| **Proposed measures to control stigma** | **60** |
| Use of visual messages for health education and sensitization | 7 |
| Survivors should use public transport to the survivor's clinic and have their transport refunded | 18 |
| Integrated management of interventions to avoid frequent visits | 12 |
| Private distribution of support items | 23 |
| **Total** | **219** |

### Thematic analysis results

#### *Thematic area I*. Type of stigma experienced by SVD survivors and their household members, Mubende and Kassanda districts, Uganda, September 2022-January 2023

SVD survivors experienced enacted, institutional, and felt stigma.

#### Enacted stigma was widely reported

All survivors interviewed, particularly those discharged during the peak of the outbreak (October 2022), experienced enacted stigma from the community in the first 2 months after discharge. There were codings related to experience of enacted stigma: [isolation and segregation by community members appeared 24 times in the 30 interviews held]. From these coded quotes, we developed the theme of “Enacted stigma based on fear from community members”. Two quotes that illustrate this theme are: *“I was treated so badly. People feared and isolated me. When I would go to the shop, they could not touch the money. [Instead] they put [out] a basket where we would drop the money and they later washed it,”* and *“They treated us very badly. The shop attendants would refuse to hold our money, [and] we were ignored by everyone. At the borehole, we always fetched water last because no one wanted to pump water after we had pumped*.*”*

Survivors discharged in the early stages of the epidemic reported that they were less likely to be stigmatized than those discharged in the later stages. Some believed that this was related to increased awareness about the severity of the disease by the community members over time.

For example, one participant commented, *“I was welcomed well by the community and I integrated well. It was still early, and the community was not yet tired of ambulances. My boss offered my job back, but I could not do it because I was weak*.*”*

Some participants mentioned the support provided to survivors to control stigma but emphasized that survivors were still considered a threat to the community, including in their own homes. For example, one participant said, *“Survivors are still stigmatized in the communities. Although people cannot openly come out to talk about it…it is implied in how they relate to them*.*”* A survivor commented, “*We are still rejected. It will take several months because now it’s been three months. Maybe after six months they will be free with us*.*”* Another said, *“To date, some people still fear and discriminate (against) us. Sometimes when you come where they are, they don’t offer you a seat, so you automatically understand they still fear you*.*”*

#### Institutional stigma was pronounced at schools, from teachers and students

Survivors further described experiencing institutional stigma, particularly at schools where survivor children or the child family members of affected households attended. Respondents reported that children from SVD-affected homes were sometimes denied access to school, while others were stopped from attending school by their parents due to rejection and verbal abuse from the teachers and fellow students. Following are two quotes demonstrating this experience: *“We received reports that some students from affected homesteads were denied end of term promotion exams…teachers fear to touch their books and fellow students also stigmatize them*.*”* and *“My children were discriminated by their friends. A time came when they did not want to go to school, so they stayed home until examination period*.*”*

#### Felt stigma: survivors feared both the response of others and that they might still be infectious

Following the intense stigma from the community, some survivors felt it was best to keep to themselves. A survivor from Mubende district said *“I have not started going to the mosque, I fear to scare people when they see me coming. It’s better not to go until…the situation is better*.*”* For some survivors, the sequelae of infection made them and others worry that they might still be infectious. One participant said, *“After discharge, I was so happy, but I feared to go home because I was still weak and thought maybe the disease was still there. I didn’t want to go home because my body was still swollen*.*”*

### Thematic area II. Causes of stigma among SVD survivors and their household members in Mubende and Kassanda districts, Uganda, September 2022–January 2023

#### Fear that survivors who looked unwell were still ill

Stigma among survivors was aggravated by the sequelae of infection and the fear of the disease. Sickly survivors were perceived as “still infectious”. One comment by a local government official describes this: *“Many of them still have health problems like scrotal swelling and pain, hearing problems, back pain mainly for the women, headache and easy irritability. The residual symptoms confirm the community’s myths that [SVD] cannot heal, further increasing the stigma among survivors*.*”* A family member also provided a related comment: *“It was hard to explain to the children why their father was still weak following discharge, so any sign like vomiting or coughing they would run away from him*.*”*

#### The extra attention to survivors created further stigma

Health organizations provided services to SVD survivors, including psychosocial support, in-kind household items and cash. While survivors appreciate the support, they and their household members also felt that this provided additional attention to them that aggravated both felt and enacted stigma. The frequent community visits by health organizations, who traveled to their community in branded vehicles, raised the attention of the community, as did the biweekly pickups from the community to the survivor’s clinic, which sometimes occurred in an ambulance. Two quotes that illustrate this experience are: *The use of ambulances to pick survivors for review [makes] people in the community think they are still sick. It is making it hard for the community to accept them as survivors*.*”* and *“Visiting their homes with so many cars, the neighbors wonder if somebody has become sick again. The frequent visits, frequent clinic reviews, the special attention with support, there is some level of stigma*.*”*

#### Public monetary and in-kind support for survivors caused further separation from community

Public provision of in-kind support was also reported to aggravate stigma for the survivors. One local government official said, *“Giving them material and monetary support in public…*., *the community segregates them because they assume they have more than other members*.*”*

#### Risk communication about survivors was confusing

Community members reported that health education and community sensitization included information that was confusing to them. For example, they reported that the messaging that survivors were safe but that they also had virus in their body fluids for up to 1 year after discharge was unclear, and that it made them uncertain about how to treat survivors. One participant said, *“A lot of stigma which was promoted during health education…. the message that these are survivors but they still have a risk, they still have the virus in their semen and tears*.*”* Furthermore, the mode of sensitization was considered ineffective to some members of the community. The vehicles promoting health education through loudspeakers were not always audible or well understood. One comment from a local government official relates to the mode of communication, *“I can remember the education I received as a youth about prevention of HIV because they were showed in video form in the community squares, but I cannot remember all that was said a few months back when the Ebola treatment unit was opened*.*”*

### Thematic area III. Effects of stigma on the SVD survivors and their household members in Mubende and Kassanda districts, Uganda, September 2022-January 2023

#### Economic effects

Survivors and their household members were affected in various ways due to the stigma experienced. Among the effects were economic interferences; a household member in Mubende shared that: *“The herdsman was not allowed to cross the compound to take the cows for grazing so he ran away and the cows died,”* while a key informant from Mubende shared: *“Most of them (the survivors) have lost their jobs. For the few that maintained their jobs, their physical health is limiting their engagement in day-to-day activities*.*”*

#### Effects on education

Additionally, stigma affected the education of school going household members to EVD survivors. Two comments by a district official and a household member describe this: *“we received reports that some students from affected homesteads were denied end of term promotion exams, teachers fear to touch their books and fellow students also stigmatize them*.*” “My children were discriminated by their friends; a time came when they did not want to go to school so they stayed home until examination period and so my children did not perform well this last term*.*”*

#### Social effects

Some families reportedly broke down due to the stigma. In some polygamous families where only one family was affected, the heads of family have refused to go back and denied them support. Other families lost the primary parent, resulting in child-headed homes or situations where children were sent to stay with their grandparents. For such homes, the subcounty in Mubende has tried to offer some support. A key informant in Mubende District shared that, *“Some families are now headed by children; the father died, the mother divorced and has refused to return in fear of getting infected”*. Furthermore, there was reportedly an increase in marital conflict caused by the close interaction among survivors. A k*ey informant in Kassanda District* shared that *“Survivors…have maintained the friendship to help console each other in case of any form of abuse. However, this close interaction among survivors has resulted in marital conflicts as wives and husbands are closer to fellow survivors than their spouses*.*”*

### Thematic area III: Proposed measures to control stigma among SVD survivors and their household members in Mubende and Kassanda districts, 2022

Respondents proposed a number of measures needed to control stigma. Among them was: use of visual messages for health education and sensitization in the communities and to intensify efforts to reintegrate survivors back to the community. A key informant from Kassanda District suggested: *“Intensify the integration of survivors in the community. We need to show that they are as normal as us and reduce on the extra attention”. Another key informant from Mubende District shared that; “Allow survivors use public means to the survivors clinic and have their transport refunded, this will reduce on the attention during pickups and drops from the clinic which causes extra attention”*. Additional measures suggested to control stigma included supporting survivors in management of their long Ebola signs and symptoms and integrated management of interventions to avoid frequent visits to survivors and their households in the community by different teams. It was suggested that all teams should move to the field together to avoid frequent visits and the so many cars that park in survivors’ compounds.

*Furthermore, r*espondents shared the need for additional support for improved wellbeing of survivors and their household members. These included a need for startup capital to restart their livelihoods, school fees for at least 6 months for their children while they worked to regain financial stability, and replacement of their phones, which were destroyed in the Ebola treatment unit as part of the response.

## Discussion

Survivors and household members of survivors in the 2022 SUDV outbreak in Uganda faced enacted, institutional, and felt stigma that existed despite attempts to reduce its impact. The stigma was largely caused by fear of ongoing infection. However, it was aggravated by prolonged SVD symptoms and some of the response efforts designed to support survivors.

SVD survivors and their household members faced enacted, institutional, and felt stigma, with enacted stigma being the most common. This finding is similar to those from other studies in Liberia and Sierra Leone, which found that EBOD survivors encountered primarily enacted and perceived external stigma, rather than internalized stigma [16, 24-26]. However, a study in Sierra Leone reported higher levels of felt stigma than enacted stigma, with social isolation having the highest impact [9]. In both the Sierra Leone study and our study, enacted stigma took the form of social isolation and discrimination for both the survivors and their household members, including children [9, 27, 28].

The prolonged SVD symptoms that persisted for survivors after discharge were seen as exacerbating the stigmatization. Persistent health problems are well-recognized among EBOD survivors, with the most common problems being partial loss of vision, dizziness, headache, sleeplessness, and myalgia [16, 29, 30]. In our evaluation, the community feared that the persistent symptoms represented potential ongoing contagiousness. This was compounded by the lack of clarity around risk communication messages passed on by healthcare workers, particularly around the possible ongoing presence of the virus in body fluids of survivors. It is important to provide clear messages to both survivors and their communities about the recovery process as well as information on what is and is not safe for survivors in language that is well-understood by the community. It may be beneficial to test different messages for understanding during inter-epidemic periods, so that such messaging is ready to deploy during outbreaks.

Response activities aimed at supporting survivors, including home visits from response staff, transportation in branded vehicles for survivors from the community to the survivor’s clinic, and offering of support items – particularly in public - also aggravated stigma. This finding has been observed in other settings, and scholars have suggested supporting the entire community as opposed to supporting only survivors as a way of minimizing the possibility for new discrimination [31]. In our study, some health education messages intended to sensitize the community on Ebola prevention measures further aggravated stigma. Similar findings have been obtained in a study of causes and consequences of EBOD outbreak stigma among children showed that children perceived specific interventions initiated to contain the epidemic, such as the ‘no touch’ policy, as primary contributors to stigma [27]. Repackaging health education messages to the community for appropriateness, integrated response interventions by partners, and private distribution of support items could help control stigma.

The stigmatization of survivors affected them economically as well as socially, through loss of work. This underscores a need to support survivors with material items during the early phases of discharge before they stabilize their economic situation; support items could include clothing, food stuffs and cash [16, 32]. In Uganda, various partners did provide both monetary and material support; however, some survivors suggested that this support was not sustainable, and that provision of monitored startup capital for small businesses for sustained self-reliance would be more effective.

Members of affected households, especially children, were also stigmatized, consistent with other studies on this topic [27, 33]. Specifically, some children were denied end-of-term exams when teachers were afraid to touch their books, while in Sierra Leone, children had ropes tied around their homes preventing them from socializing and attending schools out of fear of ongoing contagion [27]. Engagement of various stakeholders, specifically in educational institutions and in the community, could help reduce stigma among children affected by EBOD. Furthermore, supporting the children’s guardians with school fees for a few months as they regain financial stability could help ensure continuity of education for children in EBOD-affected homesteads.

Support provided to EVD survivors to attempt to reduce the impact of stigma, which included psychosocial support and community engagements to improve acceptance, appeared to be insufficient, as stigma persisted for survivors. Similar findings have been noted in other studies, where survivors emphasized the critical need for comprehensive discharge counseling as well as facilitation of reentry into the community by professional psychosocial support counselors [16]. However, other researchers have reported better coping for EVD survivors following similar support by family, friends, and prayer groups rather than professional counselors [26, 34].

Additional studies conducted in Uganda would help contextualize the required support and approaches for application in future outbreaks.

Taken together, these results suggest a need for clearer and more effective messages to both survivors and their communities about the recovery process and safety, yet also that this should be done in a way that doesn’t escalate attention towards survivors and increase stigma. The means for achieving this could be best explored during periods in between outbreaks. Also, economic supports, both for survivor children (e.g., school fees), and adults (small business startup support) should be considered as ways of increasing social stability and an acceptance for EBOD survivors.

### Study limitations

Because our sampling approach was based on convenience and willingness to participate, there may have been bias in who entered the study; persons who were home may have differed from those who were not. Household members who did not consent (n=2) may have had different experiences than those who did consent (n=10). Furthermore, household members of SVD patients who died were not included, and thus we do not know if their experience differed from those in which the case survived.

## Conclusion

Survivors experienced felt stigma, enacted stigma, and structural stigma that persisted even after implementation of control measures. The stigma was aggravated by the prolonged SVD symptoms and outbreak response and control activities such as additional attention to survivors from responders (including home visits by health workers, public distribution of support items, and conspicuous transport from home to the survivor’s clinic) and unclear health education messages about survivors. The stigma affected the education of school going household members and caused economic and social disruptions. Strengthening community engagement to counteract stigma, rethinking response activities that aggravate stigma, management of long-term SUDV symptoms for survivors, integrated response interventions by partners, private distribution of support items, and increasing awareness and sensitization could reduce stigma among the SVD survivors.

## Data Availability

The datasets upon which our findings are based belong to the Uganda Public Health Fellowship Program. For confidentiality reasons the datasets are not publicly available. However, the data sets can be availed upon reasonable request from the corresponding author and with permission from the Uganda Public Health Fellowship Program.

## List of Abbreviations

SVD: Sudan virus disease
ETU: Ebola treatment unit
VHT: Village Health teams
IDI: In-depth interview
EVD: Ebola virus disease
MoH: Ministry of Health
CFR: Case Fatality Rate
CDC: Centers for Disease Control and Prevention

## Declarations

### Ethical considerations

Before starting the project, a non-research determination form was submitted to the US Centers for Disease Control and Prevention (CDC) as a requirement. The Office of the Associate Director for Science at the CDC determined that the project did not involve human subjects research. This determination was made because the project aimed to address a public health problem and had the primary intent of public health practice. Further administrative approval to conduct this study was obtained from Mubende and Kassanda District offices, Mubende regional referral hospital case management team, the National Institute of Public Health, and Uganda Ministry of Health. Before data collection, written informed consent was sought from respondents, they were informed that their participation was voluntary and their refusal would not result in any negative consequences. To protect the confidentiality of the respondents, each was assigned a unique identifier which was used instead of their names. This activity was reviewed by CDC and was conducted consistent with applicable federal law and CDC policy.

§ §See e.g., 45 C.F.R. part 46, 21 C.F.R. part 56; 42 U.S.C. §241(d); 5 U.S.C. §552a; 44 U.S.C. §3501 et seq.

## Consent for publication

Not applicable.

## Funding and Disclaimer

This project was supported by the President’s Emergency Plan for AIDS Relief (PEPFAR) through the US Centers for Disease Control and Prevention Cooperative Agreement number GH001353 through Makerere University School of Public Health. Its contents are solely the responsibility of the authors and do not necessarily represent the official views of the US Centers for Disease Control and Prevention, the Department of Health and Human Services, Makerere University School of Public Health, or the MoH. The staff of the funding body provided technical guidance in the design of the study, ethical clearance and collection, analysis, and interpretation of data, and in writing the manuscript.

## Authors’ Contributions

GMZ, BS, ZK, JFZ, PCK, MWW, PK, SNK, HNN, BA and RZ collected data under technical guidance and supervision of JH, ARA, DK, RM, BK, JG, SP, ERG, EJN, AA and JK. GMZ analyzed and interpreted the data. GMZ drafted the manuscript. GMZ, LB, ARA, JH critically reviewed the manuscript for intellectual content. All co-authors read and approved the final manuscript.

GMZ is the guarantor of the paper.

## Acknowledgements

We acknowledge the Ministry of Health Uganda, members of the Kassanda and Mubende district health teams, Baylor Uganda and the village health teams for their Support. We further appreciate the CHWs for both Kassanda and Mubende for the guidance in the community during data collection.

## References

1. Jacob, S.T., et al., Ebola virus disease. Nature reviews Disease primers, 2020. 6(1): p. 1–31.

2. Lefebvre, A., et al., Case fatality rates of Ebola virus diseases: a meta-analysis of World Health Organization data. Médecine et maladies infectieuses, 2014. 44(9): p. 412–416.

3. Kadanali, A. and G. Karagoz, An overview of Ebola virus disease. Northern clinics of Istanbul, 2015. 2(1): p. 81.

4. Muyembe-Tamfum, J.-J., et al., Ebola virus outbreaks in Africa: past and present. Onderstepoort Journal of Veterinary Research, 2012. 79(2): p. 06–13.

5. Kinsman, J., “A time of fear”: local, national, and international responses to a large Ebola outbreak in Uganda. Globalization and health, 2012. 8(1): p. 1–12.

6. Organisation, W.H., Ebola Virus Disease. 2021. p. https://www.who.int/news-room/fact-sheets/detail/ebola-virus-disease?

7. Ibrahim, S.K., et al., Sudan virus disease outbreak in Uganda: urgent research gaps. BMJ Global Health, 2022. 7(12): p. e010982.

8. WHO, Uganda declares end of Ebola disease outbreak. 2023.

9. James, P.B., et al., An assessment of Ebola-related stigma and its association with informal healthcare utilisation among Ebola survivors in Sierra Leone: a cross-sectional study. BMC Public Health, 2020. 20(1): p. 1–12.

10. Acharibasam, J.W., B. Chireh, and H.G. Menegesha, Assessing anxiety, depression and insomnia symptoms among Ebola survivors in Africa: A meta-analysis. PLoS One, 2021. 16(2): p. e0246515.

11. Yadav, S. and G. Rawal, The current mental health status of Ebola survivors in Western Africa. Journal of clinical and diagnostic research: JCDR, 2015. 9(10): p. LA01.

12. Control, C.f.D. and Prevention, 2014 Ebola Outbreak in West Africa Epidemic Curves. 2017.

13. Bah, A.J., et al., Prevalence of anxiety, depression and post-traumatic stress disorder among Ebola survivors in northern Sierra Leone: a cross-sectional study. BMC public health, 2020. 20(1): p. 1–13.

14. Dovidio, J.F., B. Major, and J. Crocker, Stigma: Introduction and overview. 2000.

15. Karamouzian, M. and C. Hategekimana, Ebola treatment and prevention are not the only battles: understanding Ebola-related fear and stigma. International journal of health policy and management, 2015. 4(1): p. 55.

16. Lee-Kwan, S.H., et al., Support services for survivors of Ebola virus disease—Sierra Leone, 2014. Morbidity and Mortality Weekly Report, 2014. 63(50): p. 1205.

17. Marais, F., et al., A community-engaged infection prevention and control approach to Ebola. Health promotion international, 2015. 31(2): p. 440–449.

18. Development, U.S.A.f.I., Restoring Hope and Dignity of Ebola Survivors in Uganda: 4 ways USAID support helped them bounce back in life. 2022: Kampala.

19. UNICEF, UNICEF UGANDA: Ebola Virus Diseas (EVD) Update. 2022.

20. Saunders, B., et al., Saturation in qualitative research: exploring its conceptualization and operationalization. Quality & quantity, 2018. 52: p. 1893–1907.

21. Fusch Ph D P.I. and L.R. Ness, Are we there yet? Data saturation in qualitative research. 2015.

22. Aspers, P., Empirical phenomenology: A qualitative research approach (The Cologne Seminars). Indo-pacific journal of phenomenology, 2009. 9(2): p. 1–12.

23. Byrne, D., A worked example of Braun and Clarke’s approach to reflexive thematic analysis. Quality & quantity, 2022. 56(3): p. 1391–1412.

24. Overholt, L., et al., Stigma and Ebola survivorship in Liberia: results from a longitudinal cohort study. PLoS One, 2018. 13(11): p. e0206595.

25. Davidson, M.C., et al., A post-outbreak assessment of exposure proximity and Ebola virus disease-related stigma among community members in Kono District, Sierra Leone: A cross-sectional study. SSM-Mental Health, 2022. 2: p. 100064.

26. Karafillakis, E., et al., ‘Once there is life, there is hope’: Ebola survivors’ experiences behaviours and attitudes in Sierra Leone, 2015. BMJ global health, 2016. 1(3): p. e000108.

27. Denis-Ramirez, E., K.H. Sørensen, and M. Skovdal, In the midst of a ‘perfect storm’: Unpacking the causes and consequences of Ebola-related stigma for children orphaned by Ebola in Sierra Leone. Children and Youth Services Review, 2017. 73: p. 445–453.

28. Nuriddin, A., et al., Trust, fear, stigma and disruptions: community perceptions and experiences during periods of low but ongoing transmission of Ebola virus disease in Sierra Leone, 2015. BMJ global health, 2018. 3(2): p. e000410.

29. Delamou, A., et al., Profile and reintegration experience of Ebola survivors in Guinea: a cross-sectional study. Tropical Medicine & International Health, 2017. 22(3): p. 254–260.

30. de St. Maurice, A., et al. Care of Ebola survivors and factors associated with clinical sequelae— Monrovia Liberia. In Open Forum Infectious Diseases. 2018. Oxford University Press US.

31. Tsakiridis, A., Reintegrating the “Other” Challenges of Stigmatization in Policies and Practice: The Case of Ebola Survivors and Their Relatives During the 2014–2016 Epidemic. Disaster studies: Exploring intersectionalities in disaster discourse, 2020: p. 187–211.

32. Mayrhuber, E.A.-S., T. Niederkrotenthaler, and R. Kutalek, “We are survivors and not a virus:” Content analysis of media reporting on Ebola survivors in Liberia. PLoS Neglected Tropical Diseases, 2017. 11(8): p. e0005845.

33. Crea, T.M., et al., Social distancing, community stigma, and implications for psychological distress in the aftermath of Ebola virus disease. Plos one, 2022. 17(11): p. e0276790.

34. Rabelo, I., et al., Psychological Distress among Ebola Survivors Discharged from an Ebola Treatment Unit in Monrovia, Liberia—A Qualitative Study. Front Public Health. 2016; 4: 142. A detailed presentation of the psychological and social consequences for Ebola virus disease survivors who were treated and discharged from an ETU, 2016.

